# Nocturnal synchronization between hippocampal ripples and cortical delta power is a biomarker of hippocampal epileptogenicity

**DOI:** 10.1101/2024.06.05.24308489

**Authors:** Takamitsu Iwata, Takufumi Yanagisawa, Ryohei Fukuma, Yuji Ikegaya, Satoru Oshino, Naoki Tani, Hui M. Khoo, Hidenori Sugano, Yasushi Iimura, Hiroharu Suzuki, Haruhiko Kishima

## Abstract

**Objective:** Hippocampal ripples are biomarkers of epileptogenicity in patients with epilepsy, and physiological features characterize memory function in healthy individuals. Discriminating between pathological and physiological ripples is important for identifying the epileptogenic (EP) zone; however, distinguishing them from waveforms is difficult. This study hypothesized that the nocturnal synchronization of hippocampal ripples and cortical delta power classifies EP and physiological hippocampi.

**Methods:** We enrolled 38 patients with electrodes implanted in the hippocampus or the parahippocampal gyrus between April 2014 and March 2023 at our institution. We classified 11 patients (11 hippocampi) into the EP group, who were pathologically diagnosed with hippocampal sclerosis, and five patients (six hippocampi) into the non-epileptogenic (NE) group, whose hippocampi had no epileptogenicity. Hippocampal ripples were detected using intracranial electroencephalography of the hippocampal or parahippocampal electrodes and presented as ripple rates per second. Cortical delta power (0.5–4 Hz) was assessed using cortical electrodes. The Pearson correlation coefficient between the ripple rates and the cortical delta power (CRD) was calculated for the intracranial electroencephalographic signals obtained every night during the recordings.

**Results:** Hippocampal ripples detected from continuous recording for approximately 10 days demonstrated similar frequency characteristics between the EP and NE groups. However, CRDs in the EP group (mean [standard deviation]: 0.20 [0.049]) were significantly lower than those in the NE group (0.67 [0.070], *F* (1,124) = 29.6, *p* < 0.0001 (group), *F* (9,124) = 1.0, *p* = 0.43 (day); two-way analysis of variance). Based on the minimum CRDs during the 10-day recordings, the two groups were classified with 94.1% accuracy.

**Conclusion:** CRD is a biomarker of hippocampal epileptogenicity.

**Key Points:** The correlation between hippocampal ripple rate and cortical delta power was evaluated for approximately 10 days in patients with drug-resistant epilepsy.

Mean correlation coefficients were significantly lower in the epileptogenic group than in the non-epileptogenic group.

The minimum value of the correlation coefficients predicts hippocampal sclerosis.

## Introduction

Identifying pathological and physiological activities using intracranial electroencephalography (iEEG) is crucial for determining epileptogenic (EP) zones. High-frequency oscillations (HFOs), neural activities occurring at frequencies above 80 Hz on iEEG, are biomarkers for epilepsy sources or pathological tissues (González Otárula et al., 2019; Kucewicz et al., 2014; Liu et al., 2016; Scott et al., 2020). HFOs have been widely used to identify EP zones and predict surgical outcomes and seizure prognosis (González Otárula et al., 2019; Jacobs et al., 2010; Jiruska et al., 2010; Liu et al., 2016; Scott et al., 2020; Zijlmans et al., 2012). They are divided into two categories based on their frequency: ripples (80–250 Hz) and fast ripples (250–600 Hz) (Engel Jr et al., 2009; Kucewicz et al., 2014). Although fast ripples are generally considered to have high pathological significance among HFOs (Jiruska et al., 2010; Yamamoto et al., 2021), they have low signal-to-noise ratios. Conversely, ripples overlap with physiological ripples in the high-gamma range (80–150 Hz), which are activated during various physiological tasks (Miller, 2019; Yamamoto et al., 2021). Therefore, discriminating between physiological and pathological HFOs is necessary to evaluate the iEEG signals of patients with epilepsy precisely; however, this discrimination is difficult (Liu et al., 2022).

In medial temporal lobe epilepsy, HFOs are linked to hippocampal sclerosis (Agudelo Valencia et al., 2021; Řehulka et al., 2019). More HFOs are detected via intraoperative EEG from the hippocampus in patients with hippocampal sclerosis than in patients with other epileptic conditions (Agudelo Valencia et al., 2021). Additionally, the non-epileptic hippocampi can be differentiated by a lower ripple rate than the epileptic hippocampi (Řehulka et al., 2019). In the hippocampus, ripples occur not only through pathology but also through physiological activity. Sharp-wave ripples (SWRs), which are ripple waves accompanied by sharp waves, are associated with cognitive functions, including memory consolidation, recall, future planning, and instructive thoughts (Buzsáki, 2015; Girardeau et al., 2009; Iwata et al., 2024; Joo and Frank, 2018; Leonard et al., 2015; Norman et al., 2019). SWR originates from synchronous neuronal firing in the CA1 region of the mammalian hippocampus (Buzsáki, 2015; Liu et al., 2022). Inhibition of the SWR causes memory impairment, and hippocampal damage impairs recall and future planning (Girardeau et al., 2009; Kurczek et al., 2015; Lah and Miller, 2008; Scoville and Milner, 1957). A hypothesis suggests that the crosstalk between physiological and pathological activities in a similar frequency range may be associated with memory loss and cognitive impairment in long-term epilepsy (Agudelo Valencia et al., 2021; Řehulka et al., 2019). Several studies have demonstrated that hippocampal interictal epileptic discharges disrupt memory and cognition (Kleen et al., 2013, 2010). Conversely, SWRs have physiological functions even with pathological ripples (Ewell et al., 2019). The precise characterization of iEEG signals in the ripple range will contribute to identifying the pathological and physiological characteristics of the hippocampus (Liu et al., 2022).

In addition to its waveform characteristics, SWR has various physiological characteristics. Although the frequency of SWRs fluctuates depending on the cognitive state while awake (Iwata et al., 2024), they exhibit diurnal fluctuations in synchronization with the sleep and wake cycles, characterized by delta power in cortical activity. Notably, SWR frequency strongly correlates with the delta power during sleep (Norman et al., 2019). However, diurnal fluctuations in the frequency of pathological ripples and their synchronization with the delta power have not yet been elucidated. This study aimed to test the hypothesis that the nocturnal synchronization of hippocampal ripples and cortical delta power decreases when the SWR is contaminated by pathological ripples in the pathological hippocampus.

## Results

### Patient groups

Among the 42 patients implanted with electrodes in the hippocampus between April 2014 and April 2023 at Osaka University Hospital, 11 patients were diagnosed with hippocampal sclerosis after hippocampal removal and were classified into the EP group (Fig. 1). The NE group comprised two patients in whom the hippocampus was resected without pathological abnormalities found, and three patients who achieved seizure-free status after resection of regions other than the hippocampus. Characteristics of the EP and NE groups were compared. No significant differences were observed in age, sex, electrode implantation site, region of implantation, electrode type, age at initial seizure, disease duration, pre- or post-operative WAIS or WMSR scores (*p* > 0.05, Wilcoxon rank-sum test [continuous variables] or chi-square test [categorical variables]) (Table 1).

**Figure 1.**
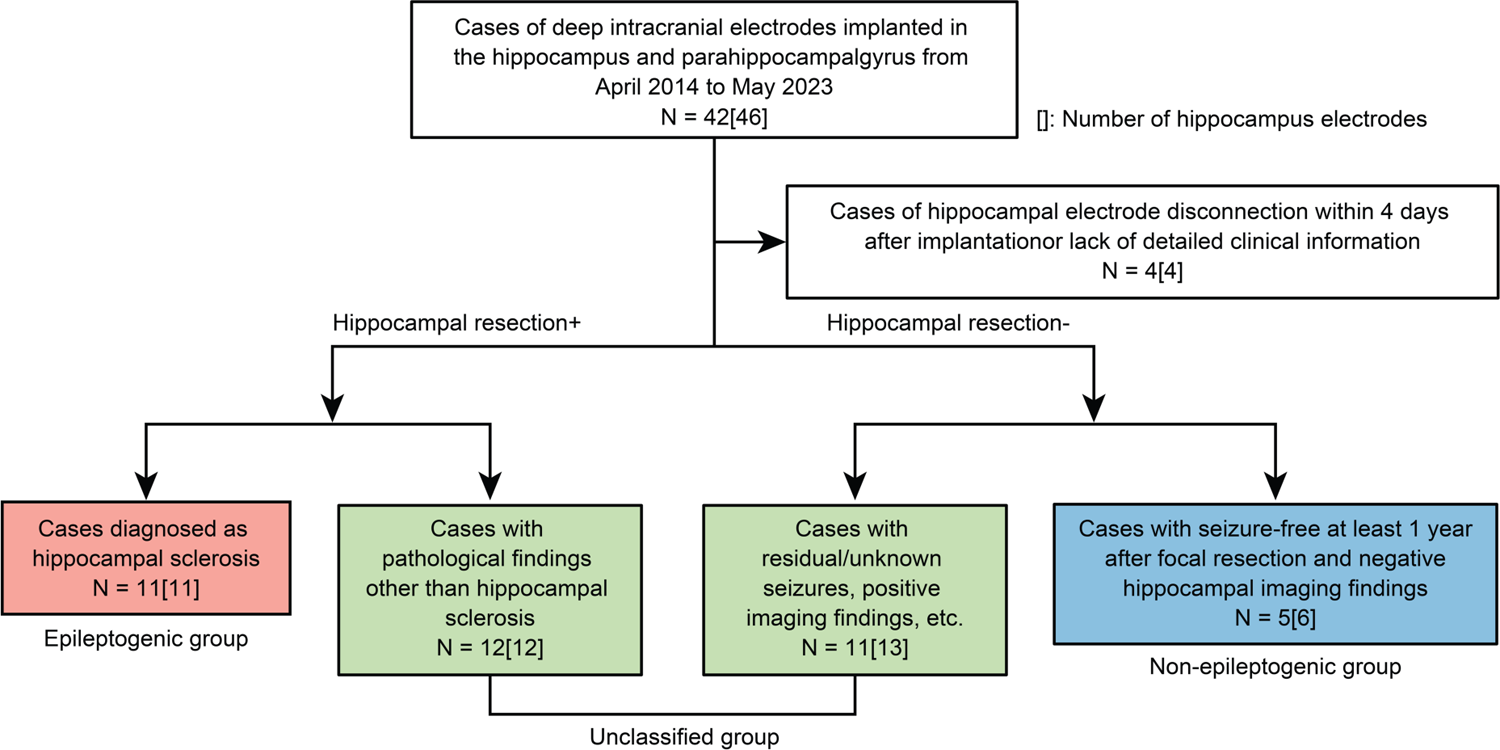
CONSORT diagram. We recorded the iEEG data of 42 patients from 2014 to 2023. The patients were classified into three groups: epileptogenic (EP) non-epileptogenic (NE), and unclassified (UC).

**Table 1.**
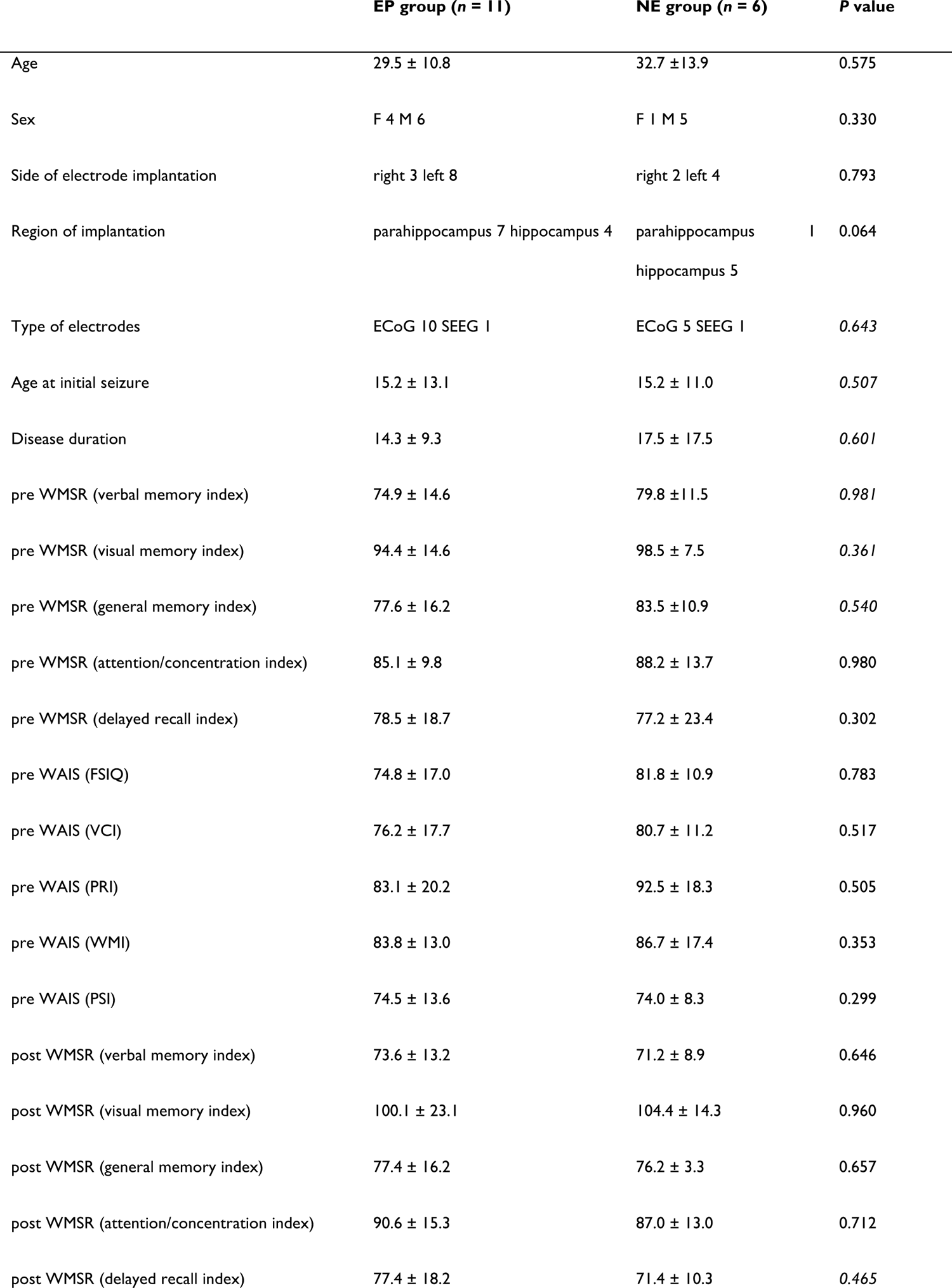
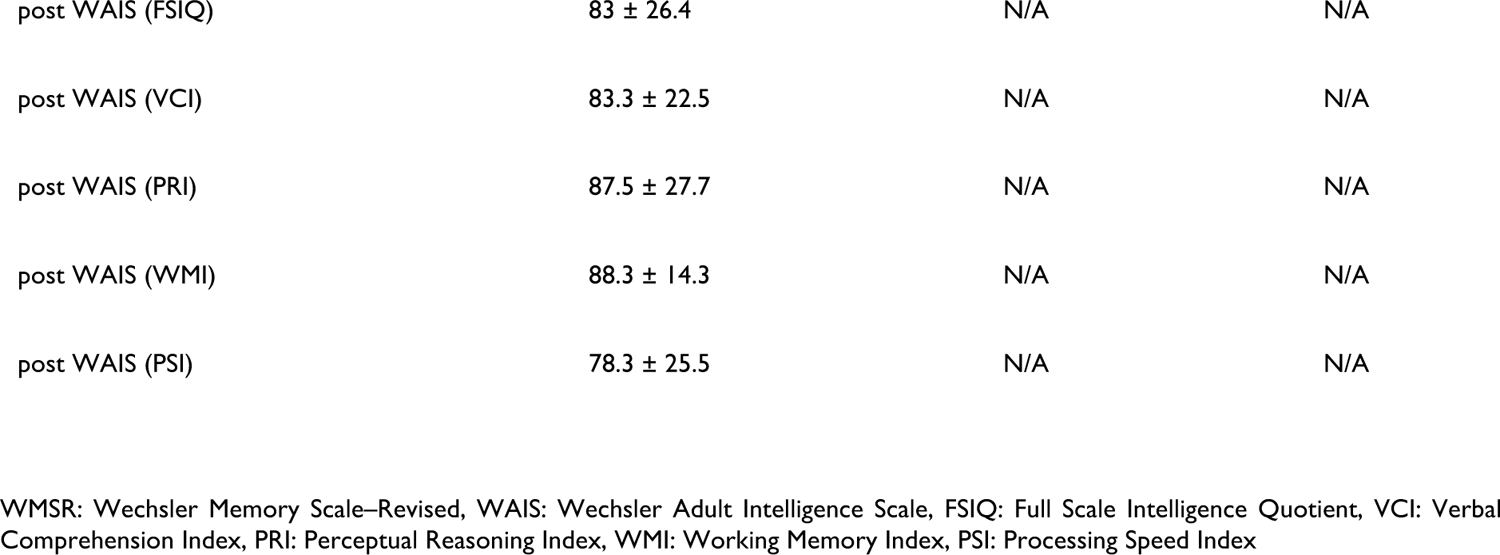
Comparison of patient characteristics between the EP and NE groups.

|  | EP group (n = 11) | NE group (n = 6) | P value |
| --- | --- | --- | --- |
| Age | 29.5 ± 10.8 | 32.7 ± 13.9 | 0.575 |
| Sex | F 4 M 6 | F 1 M 5 | 0.330 |
| Side of electrode implantation | right 3 left 8 | right 2 left 4 | 0.793 |
| Region of implantation | parahippocampus 7 hippocampus 4 | parahippocampus 1<br>hippocampus 5 | 0.064 |
| Type of electrodes | ECoG 10 SEEG 1 | ECoG 5 SEEG 1 | 0.643 |
| Age at initial seizure | 15.2 ± 13.1 | 15.2 ± 11.0 | 0.507 |
| Disease duration | 14.3 ± 9.3 | 17.5 ± 17.5 | 0.601 |
| pre WMSR (verbal memory index) | 74.9 ± 14.6 | 79.8 ± 11.5 | 0.981 |
| pre WMSR (visual memory index) | 94.4 ± 14.6 | 98.5 ± 7.5 | 0.361 |
| pre WMSR (general memory index) | 77.6 ± 16.2 | 83.5 ± 10.9 | 0.540 |
| pre WMSR (attention/concentration index) | 85.1 ± 9.8 | 88.2 ± 13.7 | 0.980 |
| pre WMSR (delayed recall index) | 78.5 ± 18.7 | 77.2 ± 23.4 | 0.302 |
| pre WAIS (FSIQ) | 74.8 ± 17.0 | 81.8 ± 10.9 | 0.783 |
| pre WAIS (VCI) | 76.2 ± 17.7 | 80.7 ± 11.2 | 0.517 |
| pre WAIS (PRI) | 83.1 ± 20.2 | 92.5 ± 18.3 | 0.505 |
| pre WAIS (WMI) | 83.8 ± 13.0 | 86.7 ± 17.4 | 0.353 |
| pre WAIS (PSI) | 74.5 ± 13.6 | 74.0 ± 8.3 | 0.299 |
| post WMSR (verbal memory index) | 73.6 ± 13.2 | 71.2 ± 8.9 | 0.646 |
| post WMSR (visual memory index) | 100.1 ± 23.1 | 104.4 ± 14.3 | 0.960 |
| post WMSR (general memory index) | 77.4 ± 16.2 | 76.2 ± 3.3 | 0.657 |
| post WMSR (attention/concentration index) | 90.6 ± 15.3 | 87.0 ± 13.0 | 0.712 |
| post WMSR (delayed recall index) | 77.4 ± 18.2 | 71.4 ± 10.3 | 0.465 |
| post WAIS (FSIQ) | 83 ± 26.4 | N/A | N/A |
| post WAIS (VCI) | 83.3 ± 22.5 | N/A | N/A |
| post WAIS (PRI) | 87.5 ± 27.7 | N/A | N/A |
| post WAIS (WMI) | 88.3 ± 14.3 | N/A | N/A |
| post WAIS (PSI) | 78.3 ± 25.5 | N/A | N/A |

WMSR: Wechsler Memory Scale–Revised, WAIS: Wechsler Adult Intelligence Scale, FSIQ: Full Scale Intelligence Quotient, VCI: Verbal Comprehension Index, PRI: Perceptual Reasoning Index, WMI: Working Memory Index, PSI: Processing Speed Index

### Hippocampal ripples of representative patients in NE and EP groups

Hippocampal ripples were detected in NE and EP groups. For two representative patients in each group, the detected ripples demonstrated features similar to those of the SWR (Fig. 2A, B)(Liu et al., 2022). The average waveforms of the detected ripples showed high-frequency ripples with sharp low-frequency waves in both patients (Fig. 2C, D). The distributions of the peak slow-wave amplitude and peak frequency were similar between the two patients (Fig. 2E, F). Based on the waveforms and frequency properties, the detected hippocampal ripples were similar in both patients.

**Figure 2.**
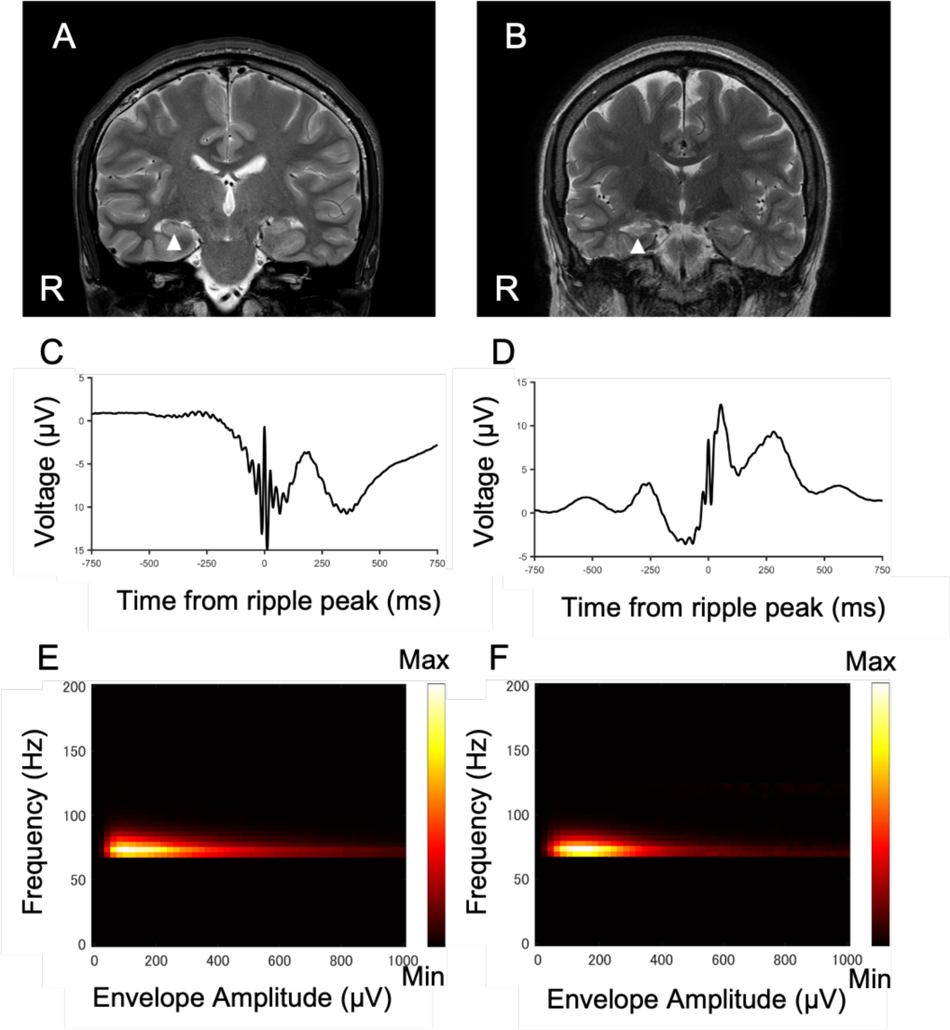
Representative hippocampal ripples detected from patients of NE and EP groups (A, B) Preoperative magnetic resonance imaging (MRI) of two representative patients; Pt 15 from NE group (A) and Pt 22 from EP group (B). The arrowhead indicates the hippocampi in which some intracranial electrodes were implanted. Right hippocampus of Pt 22 shows hippocampal sclerosis. (C, D) Mean waveform of field potential recorded from the implanted electrodes of patients from NE group (C) and EP group (D). (E, F) The distribution of peak slow wave amplitude and peak frequency of every ripple event was shown as color map for the representative patients from NE group (E) and EP group (F). Density plot reveals that all events were included in a single cluster of similar frequency and envelope amplitude for both patients.

### Diurnal fluctuations in hippocampal ripples

Hippocampal ripple rate and cortical delta power were assessed using long-term iEEG recordings (Fig. 3A and B). For representative patients in both the NE and EP groups, cortical delta power increased during the night and decreased during the daytime, indicating diurnal fluctuations (*p* < 0.0001, *F*_143,1034_ = 3.66, *n* = 1178 time points (NE group); *p* < 0.0001, *F*_143,997_ = 4.12, *n* = 1215 time points (EP group); one-way ANOVA). Although antiepileptic agents were reduced during the long-term recording, consistent diurnal fluctuations in delta power were observed throughout (Supplementary Fig. 2). Additionally, the hippocampal ripple rates of patients in the NE group were synchronized with delta powers, with high CRDs occurring daily (correlation coefficient: 0.773 [0.731–0.786]). However, the CRD decreased on the day of the seizure. The averages of the ripple rates and delta power over 9 days exhibited synchronized diurnal fluctuations in the ripple rate and delta power (Fig. 3C). Conversely, for a patient in the EP group, the ripple rate increased during the daytime and decreased at night, indicating an inverse correlation with delta power (Fig. 3D, −0.419 [−0.461 to −0.342]). Additionally, the CRD further decreased during the withdrawal period of antiepileptic agents with seizures. The minimum CRD for each day differed between two representative patients in the EP (−0.587) and NE (0.406) groups.

**Figure 3.**
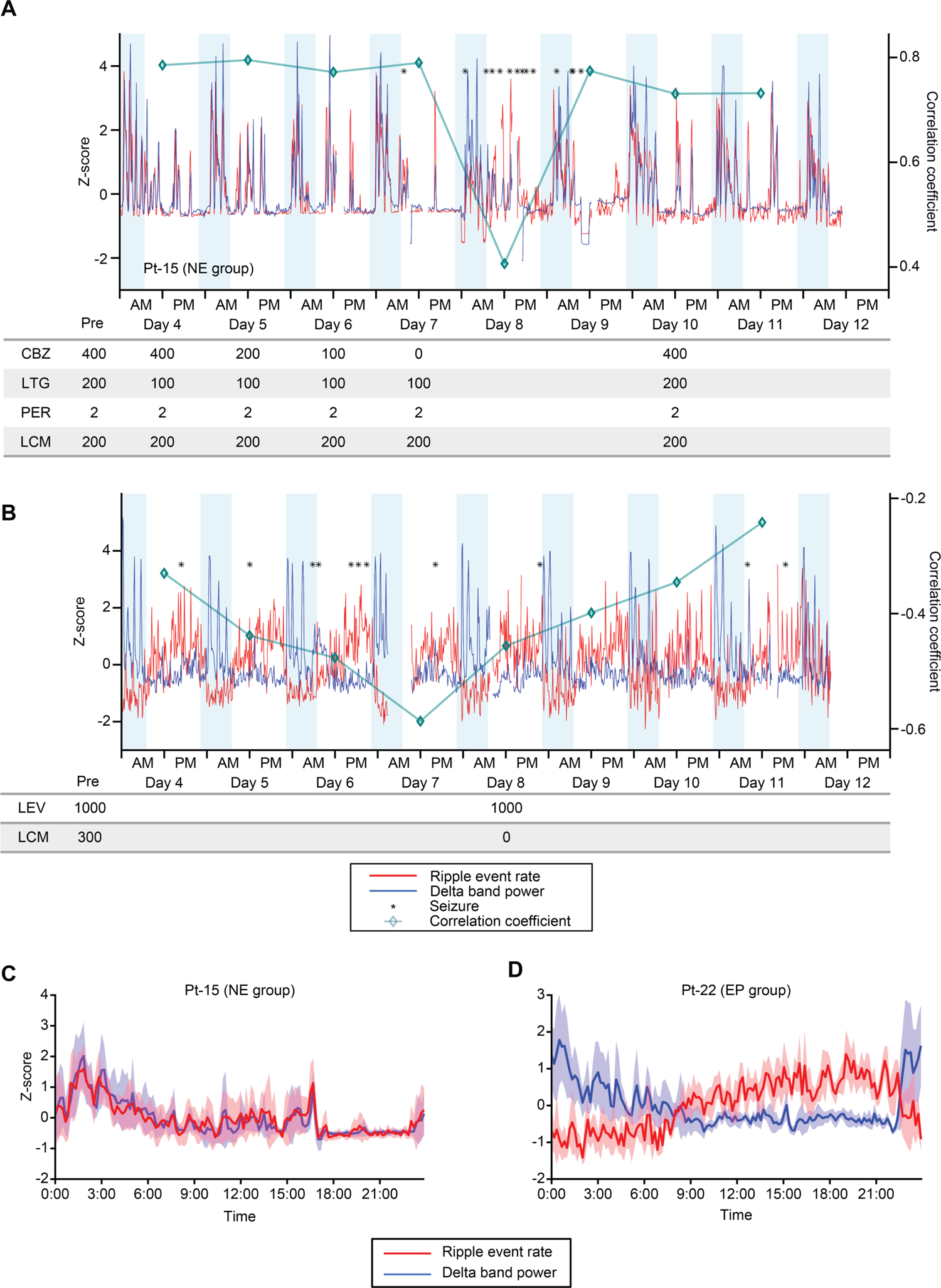
Representative examples of the hippocampal ripple event rates, cortical delta power, and seizures during 10 days of iEEG recording. (**A**) A time course of the hippocampal ripple event rates (red) and cortical delta power (blue) for nine consecutive days in a representative patient in the NE group (Pt-15). The light blue background indicates the time of day when the room was darkened. The square symbol represents the correlation coefficient between the ripple event rate and the delta power for the whole day. The asterisk indicates the timing of the seizure. The lower column indicates the amount of the antiepileptic agent. (**B**) A time course of the ripple event rates (red) and cortical delta power (blue) for nine consecutive days in a representative patient in the EP group (Pt-22). The light blue background indicates the time of day when the room was darkened. The square symbol represents the correlation coefficient between the ripple event rate and the delta power for the whole day. The asterisk indicates the timing of the seizure. The lower column indicates the amount of the antiepileptic agent. (**C, D**) The lines show the mean and 95% confidence intervals of the Z-scoring ripple event rates (red) and delta power (blue) over 24 h in patients in the NE group (**C**) and EP group (**D**).

### CRDs differed among patient groups and changed with seizure frequency

CRDs at night were compared between the two groups. The ripple event rate between 22:00 (lights out time) and 7:00 (lights on time) was analyzed. Over the 10 days of recordings among the two groups, CRD during the night significantly differed between the EP (mean (SD): 0.202 (0.049)) and NE (0.665 (0.070)) groups, although the variation in the CRD among days was not significant (*F* (1,124) = 29.6, *p* < 0.0001 (group), *F* (9,124) = 1.0, *p* = 0.4305 (day) two-way ANOVA) (Fig. 4A).

**Figure 4.**
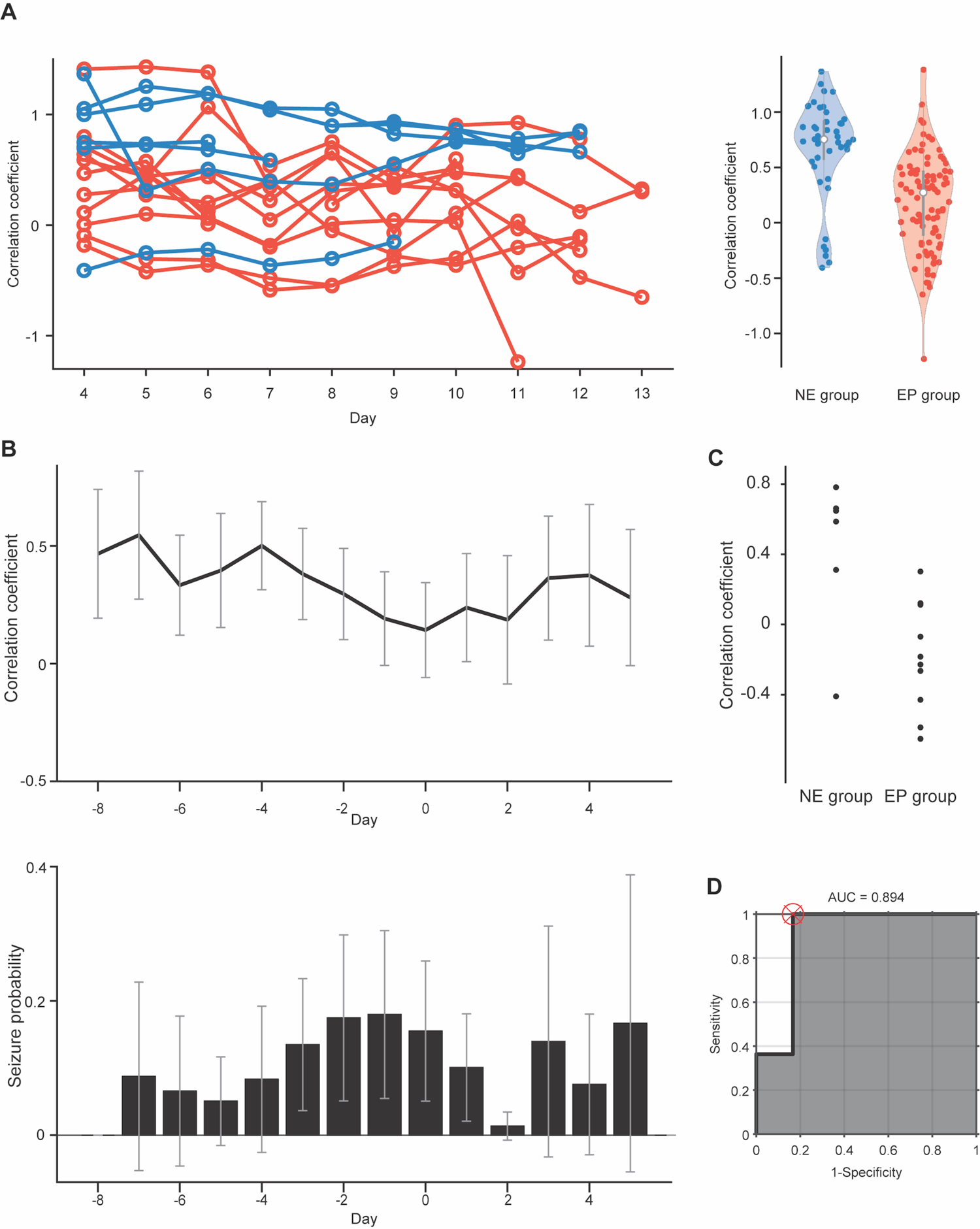
CRD of long-term recording differs between EP and NE groups. (**A**) The Fisher z-transformed correlation coefficients between the hippocampal ripple event rates and the cortical delta power are plotted against the day after electrode implantation for all patients in the EP (red) and NE (blue) groups. Violin plots of the Fisher z-transformed correlation coefficients are shown for each group. (**B**) Mean correlation coefficients aligned on the day with the minimum correlation coefficient are plotted with the mean frequency of the seizure events. The error bar shows the 95% confidence interval. (**C**) The minimum value of the correlation coefficient between the hippocampal ripple event rate and the cortical delta power for each patient is plotted for each group. (**D**) ROC curve for classifying the NE and EP groups based on the minimum value of the correlation coefficient.

Among these patients, seizure frequency increased as CRD decreased. When we averaged the CRDs of all patients by aligning the day with the minimum CRD to day 0, seizure frequency tended to increase 1–2 days before the day with the minimum CRD (Fig. 4B). Notably, the average seizure frequency and average CRD did not change in parallel among patients and were not significantly correlated (*R* = 0.0423, *p* = 0.8763).

### Detection of hippocampal sclerosis based on minimum CRDs

We examined whether the two groups could be classified based on the lowest CRD during electrode implantation. The results showed that the two groups could be classified with 94.1% accuracy (AUC: 0.894; sensitivity: 100.0%; specificity: 83.3%) when using a CRD of 0.3 as the classification criterion, as depicted in Fig. 4C and D.

## Discussion

CRD indicates hippocampal epileptogenicity. The CRDs at night were significantly lower in the EP group than in the NE group, which had hippocampal sclerosis. In this cohort, a CRD of less than 0.3 predicted hippocampal sclerosis with 94.1% accuracy. Despite similar waveforms and frequency characteristics between the NE and EP groups, nocturnal synchronization between the hippocampal ripples and cortical delta power has been proposed as a biomarker of hippocampal epileptogenicity.

Previous studies demonstrated that epileptic ripples are biomarkers of epileptogenicity in patients with epilepsy (Agudelo Valencia et al., 2021; Akiyama et al., 2011; Jacobs et al., 2010; Jiruska et al., 2010; Zijlmans et al., 2012). A greater percentage of ripples was found in the seizure onset zone than in normal tissue (González Otárula et al., 2019); thus, the EP hippocampus can be inferred based on the ripple rate in unilateral mesial temporal lobe epilepsy (Řehulka et al., 2019). However, these studies did not distinguish between pathological and physiological ripples. In this study, we hypothesized that epileptogenicity could be assessed by estimating CRD because normal hippocampal SWR strongly correlates with sleep depth, as indicated by delta power. Particularly, we assessed the relationship between epileptogenicity and CRD by comparing two patient groups, with and without hippocampal sclerosis, and increasing the epileptogenicity in each patient by reducing the use of antiepileptic agents. The CRD tended to decrease when seizure severity increased in response to reduced antiepileptic agent administration (Supplementary Fig. 4). Although the variation in CRDs during treatment with reduced antiepileptic agents was not statistically significant, seizure frequency increased 1–2 days before the day with the minimum CRDs, occurring between the hippocampal ripples and the cortical delta power. Decreasing CRDs has been suggested to indicate increased epileptogenicity. Notably, the seizure frequency was not maximal on the day with the minimum CRDs, implying that increased seizures were not a direct cause of the decreased correlation through epilepsy-related EEG changes, such as an increase in interictal EP discharges. The increase in epileptogenicity owing to the reduction in antiepileptic agents has been suggested to increase the number of pathological ripples, thereby reducing CRD.

Interestingly, the CRDs were significantly different between patients with and without hippocampal sclerosis, suggesting that epileptogenicity can be identified by a decreased CRD. For some patients in the EP group, the hippocampal ripple rate and cortical delta power were inversely correlated, suggesting that pathological ripples are not only out of phase with delta power but also in the reversed phase. Theoretically, if pathological ripples are desynchronized with cortical delta power, the CRD, which is the sum of pathological and physiological ripples, should approach zero rather than become negative as pathological ripples increase. This reversed-phase coupling between hippocampal ripples and cortical delta power may be related to hippocampal sclerosis. Moreover, an abnormal CRD may predict epileptogenicity rather than pathology and could be applicable to other hippocampal epileptic conditions, such as brain tumors and focal cortical dysplasia. Further studies on various EP lesions in the hippocampus will determine the generalizability of these findings.

Although this study demonstrated a diurnal rhythm in ripple fluctuations, previous studies have reported diurnal variations in EP activity with 24-h rhythms or even multiday rhythms (Baud et al., 2018; Karoly et al., 2018; Khan et al., 2018). Notably, increased interictal spikes have been reported during slow-wave sleep (Frauscher et al., 2015; Sammaritano et al., 1991). Additionally, interictal activity during wakefulness and REM sleep has been useful for predicting epileptogenicity (Sammaritano et al., 1991). Similarly, increased interictal epileptiform discharges during NREM sleep increases delta power, affecting cognitive function (Bölsterli et al., 2017; Halász et al., 2019). Moreover, other reports have highlighted an association between epilepsy and REM sleep, which contrasts its relationship with NREM sleep (Ikoma et al., 2023; Sadak et al., 2022; Sedigh-Sarvestani et al., 2014). Our results revealed two types of abnormal correlations within the EP group: one synchronized with the cortical delta power with decreasing correlation, and the other synchronized in the reversed phase. These differences may explain the relationship between epileptogenicity and different sleep features such as REM and NREM. Large-scale studies focusing on the synchrony between epileptic and normal EEG patterns and their relationship to various types of epilepsy are warranted.

This study has certain limitations. First, the retrospective nature of our study may have introduced bias, necessitating validations in prospective studies, despite our focus on patients with and without pathologically identified hippocampal sclerosis. Second, the sample size was relatively small, limiting the generalizability of the findings. However, the demonstration of reduced CRD in the EP hippocampus provides the rationale for conducting a larger cohort study to confirm the generalizability of our findings. Finally, we relied on invasive data acquisition techniques, which may not be feasible in all clinical settings. Distinguishing between pathological and physiological ripples is difficult, even with invasive techniques. Our results suggest that CRD during long-term recordings is a useful biomarker for identifying hippocampal epileptogenicity without classifying each hippocampal ripple.

In conclusion, this study provides a novel biomarker for assessing epileptogenicity of hippocampal epileptogenicity and evaluates the correlation between hippocampal ripples and cortical delta power. These findings may lead to improved diagnostic and treatment strategies for patients with refractory epilepsy.

## Materials &Methods

### Patients

From April 2014 to April 2023, 42 patients underwent intracranial electrode implantation in the hippocampus or parahippocampal gyrus for the focal diagnosis of refractory epilepsy. However, four patients were excluded because of disconnection of the hippocampal electrodes within 4 days of implantation (3) or lack of detailed clinical information (1) (Fig. 1). Patient characteristics are summarized in Table 2. The remaining patients were divided into two groups: those with hippocampal sclerosis and those without epileptogenicity in the hippocampus. For the EP group, we selected patients who underwent focal resection after electrode implantation, had their hippocampi resected, and were pathologically diagnosed with hippocampal sclerosis. For the non-epileptogenic (NE) group, we selected patients with no significant magnetic resonance imaging findings in the hippocampus and no seizures for at least 1 year after the resection of areas other than the hippocampus. The remaining patients were assigned to the unclassified group.

**Table 2.** Patient characteristics.

|  | Value |
| --- | --- |
| Age (years) | 28.3 ± 12.5 |
| Sex | F 14 M 24 |
| Side of electrode implantation | right 11 left 23 bilateral 4 |
| Region of implantation | parahippocampus 17 hippocampus 25 |
| Electrode implantation period (days) | 10.8 ± 2.0 |
| Type of electrodes | ECoG 29 SEEG 9 |
| Age at initial seizure (years) | 14.2 ± 10.3 |
| Disease duration (years) | 14.1 ± 9.8 |
| pre WMSR (verbal memory index) | 79.7 ± 17.4 |
| pre WMSR (visual memory index) | 95.1 ± 18.0 |
| pre WMSR (general memory index) | 81.3 ± 17.9 |
| pre WMSR (attention/concentration index) | 87.5 ± 12.8 |
| pre WMSR (delayed recall index) | 80.1 ± 16.7 |
| pre WAIS (FSIQ) | 78.9 ± 14.8 |
| pre WAIS (VCI) | 80.1 ± 16.7 |
| pre WAIS (PRI) | 87.4 ± 15.4 |
| pre WAIS (WMI) | 82.4 ± 14.6 |
| pre WAIS (PSI) | 79.7 ± 15.6 |
| post WMSR (verbal memory index) | 76.9 ± 14.7 |
| post WMSR (visual memory index) | 100.3 ± 17.1 |
| post WMSR (general memory index) | 80.1 ± 13.9 |
| post WMSR (attention/concentration index) | 89.0 ± 12.6 |
| post WMSR (delayed recall index) | 75.4 ± 15.4 |
| post WAIS (FSIQ) | 79.4 ± 17.3 |
| post WAIS (VCI) | 78.7 ± 15.5 |
| post WAIS (PRI) | 87.6 ± 20.7 |
| post WAIS (WMI) | 82.0 ± 16.4 |
| post WAIS (PSI) | 79.0 ± 19.0 |

WMSR: Wechsler Memory Scale–Revised, WAIS: Wechsler Adult Intelligence Scale, FSIQ: Full Scale Intelligence Quotient, VCI: Verbal Comprehension Index, PRI: Perceptual Reasoning Index, WMI: Working Memory Index, PSI: Processing Speed Index

We retrospectively reviewed patient characteristics such as age, sex, side of the hippocampal electrode, location of electrodes, initial seizure age, disease duration, pre- and post-surgical Wechsler Memory Scale-Revised (WMSR) scores, pre- and post-surgical Wechsler Adult Intelligence Scale (WAIS) scores, hippocampal pathological findings, and seizure outcomes. If any index of the component within the framework of the WAIS or WMSR decreased to less than 50, it was systematically adjusted to 50, given that a precise calculation of the exact value was infeasible.

### iEEG acquisition and processing

The electrode placement was determined solely based on clinical necessity. Our clinical team determined the best placement of the electrodes for all the patients required to localize the EP regions. The EEG data analyst did not participate in this process. Data were collected from the Department of Neurosurgery at Osaka University Hospital. The research protocol was approved by the Ethical Review Board Osaka University Hospital (approval no. 14353, UMIN000017900), and informed consent was obtained from all participants. Data were acquired from the participants’ rooms during their hospital stays. The analysis included only EEG data from 4 days post-implantation to minimize the impact of surgery. Based on visual inspection of the recorded EEG data, seizures and segments of intense epileptic activity were discarded from subsequent analyses.

Intracranial EEG signals were recorded from the subdural and depth electrodes at 10 kHz using an EEG-1200 instrument (Nihon Kohden, Tokyo, Japan). A low-pass filter with a cutoff frequency of 3,000 Hz and a high-pass filter with a cutoff frequency of 0.016 Hz were employed for recording, and downsampling to 2 kHz was performed using an eighth-order Chebyshev Type I low-pass filter before resampling. For the reference electrodes, we used either subgalea electrodes facing the galea or skull-penetrating electrodes, although intracranial electrodes were used in only a few patients. The subdural contacts were arranged in both grid and strip configurations with an intercontact spacing of 10 mm and a contact diameter of 3 mm. The intercontact spacing of the deep electrode contacts ranged from 5 to 15 mm, with a width of 1 mm. Electrode localization was accomplished by co-registering postoperative computed tomography data with preoperative T1-weighted or fluid-attenuated inversion recovery MRI data using SYNAPSE VINCENT (Fujifilm, Tokyo, Japan).

### Ripple detection

Hippocampal ripples were identified using a previously reported method (Liu et al., 2022; Norman et al., 2019). The local field potentials (LFPs) of the hippocampal electrodes at the selected sites were converted into bipolar signals between the adjacent electrodes (Supplementary Fig. 5A). Periods associated with epileptic activity or movement artifacts were excluded to detect the candidate ripples. To remove power-line noise at 60, 120, 180, and 240 (± 1.5) Hz, we used a 1,150-order finite impulse response (FIR) bandpass filter. Subsequently, the signals were filtered between 70 Hz and 180 Hz using a zero-lag linear-phase Hamming-window FIR filter with a transition bandwidth of 5 Hz (Supplementary Fig. 5B). The instantaneous amplitude was computed using Hilbert transformation and clipped at four times the standard deviation (SD). Finally, the clipped signal was squared and low-pass filtered to calculate the mean and SD. Ripple candidates were selected from a range in which the unclipped signal exceeded four times the SD. Ripple candidates with durations shorter than 20 ms or longer than 200 ms were rejected, and peaks closer to 30 ms were concatenated. The detected ripple rate was normalized to the mean and SD for each day.

### Time-frequency analysis of the detected ripple

The method outlined by Ewell et al. (Ewell et al., 2019) was applied to the dataset to assess the pathology of the detected ripples. The peak slow-wave amplitude and frequency were determined for each ripple event. To measure the slow wave amplitude, 500 milliseconds data segments centered on the ripple event were band-pass filtered between 0.2 Hz and 40 Hz, and the maximum absolute amplitude of the filtered data was taken. Peak frequency was calculated by applying a fast Fourier transform to the data segments. The highest power peak at frequencies greater than 70 Hz was identified as the peak frequency. The density plots of the detected peaks for the frequency and envelope amplitudes are shown.

### Correlation between the hippocampal ripples and cortical delta power

To assess the correlation between hippocampal ripples and cortical delta power, delta power (0.5–4 Hz) was evaluated via electrocorticography in patients implanted with cortical electrodes. For some patients implanted with stereotactic electrodes only, cortical delta powers were evaluated using the LFPs from the electrodes in the area penetrating the cortex. Electrodes contaminated with artifacts were visually identified and excluded from the calculations. Bipolar potentials were obtained using all the available pairs of adjacent cortical electrodes. Periods associated with epileptic activity or movement artifacts were excluded from the analysis. The power-line noise at 60, 120, 180, and 240 (±1.5) Hz was removed with a 1,150-order FIR band stop filter. To avoid edge effects, we excluded 10 s from either end of each analysis interval from the analysis. These potentials were filtered using a high-pass filter (cutoff: 0.5 Hz) and a low-pass filter (cutoff: 4 Hz), and the Hilbert transformation was applied. The Hilbert function was used to obtain the absolute value of the instantaneous amplitude of the analyzed signal. The ripple event rate and cortical delta power were calculated every 10 min. The delta power of each electrode pair was normalized and averaged. Data were normalized to the mean and SD for each day. Finally, we computed the correlation coefficient between the hippocampal ripple rates and cortical delta power (CRD) every 10 min using Pearson’s correlation coefficient. The calculated correlation coefficients were transformed using Fisher’s z-transform. These computations were performed using MATLAB 2017b.

### Classification of hippocampal sclerosis based on CRDs

To predict hippocampal pathology, we classified the participants based on the relationship between hippocampal ripples and cortical delta power. We calculated CRD at night (from 11 pm to 7 am) each day. Days with no measurements for > 12 hours were excluded from the analysis. The CRDs varied with decreasing doses of antiepileptic drugs, and the lowest point was considered to reflect the pathological state (Supplementary Table 1). We used the minimum CRD value for each participant to classify the EP and NE groups. The area under the curve (AUC) was calculated using the receiver operating characteristic (ROC) curves to assess the accuracy of the binary classification using the CRD as the threshold.

### Statistical analysis

Unless otherwise specified, significant differences were assessed using analysis of covariance (ANOVA), Wilcoxon rank-sum test, and chi-square test. Pearson’s correlation analysis was used to evaluate correlations. The exact probability value (*p-*value) was calculated as the alpha level, with *p* < 0.05 defined as significant. This study used observational data, and data collection and analysis were performed using MATLAB 2017b.

## Author contributions

**Takamitsu Iwata**: Conceptualization, data curation, formal analysis, methodology, visualization, writing–original draft preparation, writing–review, and editing. **Takufumi Yanagisawa**: Conceptualization, data curation, formal analysis, methodology, and writing– original draft preparation. **Yuji Ikegaya**: Writing, reviewing, and editing. **Ryohei Fukuma**: Methodology. **Satoru Oshino**: Data curation, writing, review, and editing. **Naoki Tani**: Data curation, writing, review, and editing. **Hui M. Khoo**: Data curation, writing, reviewing, and editing. **Hidenori Sugano**: Writing, reviewing, and editing. **Yasushi Iimura**: Writing, review, and editing. **Hiroharu Suzuki**: Writing, review, and editing. **Haruhiko Kishima**: Supervision, data curation, writing, reviewing, and editing.

## Acknowledgments

This study was supported by the Japan Science and Technology Agency (JST) Exploratory Research for Advanced Technology (JPMJER1801). This research was supported in part by the Core Research for Evolutional Science and Technology (JPMJCR18A5), Moonshot R&D (JPMJMS2012), Grants-in-Aid for Scientific Research from KAKENHI (20H05705), and Grants from the Japan Agency for Medical Research and Development (AMED) (19 dm02070h0001, 19 dm0307103, and 19 dm0307008). The authors thank R. Hioki and A. Kambara for their technical assistance.

## Data availability statement

The datasets generated and analyzed during the current study are available on Figshare at https://figshare.com/s/XXXXXXXXXXX.

## Patient consent statement

The authors confirm that written informed consent has been obtained from the involved patients or, if appropriate, from the parent, guardian, or agent with the power of attorney regarding the involved patients and that they have given approval for this information to be published.

