## Supplementary information for "Nocturnal synchronization between hippocampal ripples and cortical delta power is a biomarker of hippocampal epileptogenicity"

### Supplementary Materials for
### 'Nocturnal synchronization between hippocampal ripples and cortical delta power predicts epileptogenicity and cognitive performance'

Takamitsu Iwata,[1] Takufumi Yanagisawa,[1,2] Yuji Ikegaya,[3,4,5] Ryohei Fukuma,[1] Satoru Oshino,[1] Naoki Tani,[1] Hui M. Khoo,[1] Hidenori Sugano,[6] Yasushi Iimura, [6] Hiroharu Suzuki, [6] and Haruhiko Kishima[1]

[1]Department of Neurosurgery, Graduate School of Medicine, Osaka University, Suita, Osaka, 565-0871, Japan
[2]Institute for Advanced Co-Creation Studies, Osaka University, Suita, Osaka, 565-0871, Japan
[3]Laboratory of Chemical Pharmacology, Graduate School of Pharmaceutical Sciences, The University of Tokyo, Tokyo 113-0033, Japan
[4]Institute for AI and Beyond, The University of Tokyo, Tokyo 113-0033, Japan
[5]National Institute of Information and Communications Technology, Center for Information and Neural Networks, Suita, Osaka, 565-0871, Japan

#### This PDF file includes the following:

Supplementary Figs. 1 to 5
Supplementary Table 1

### Figure 1

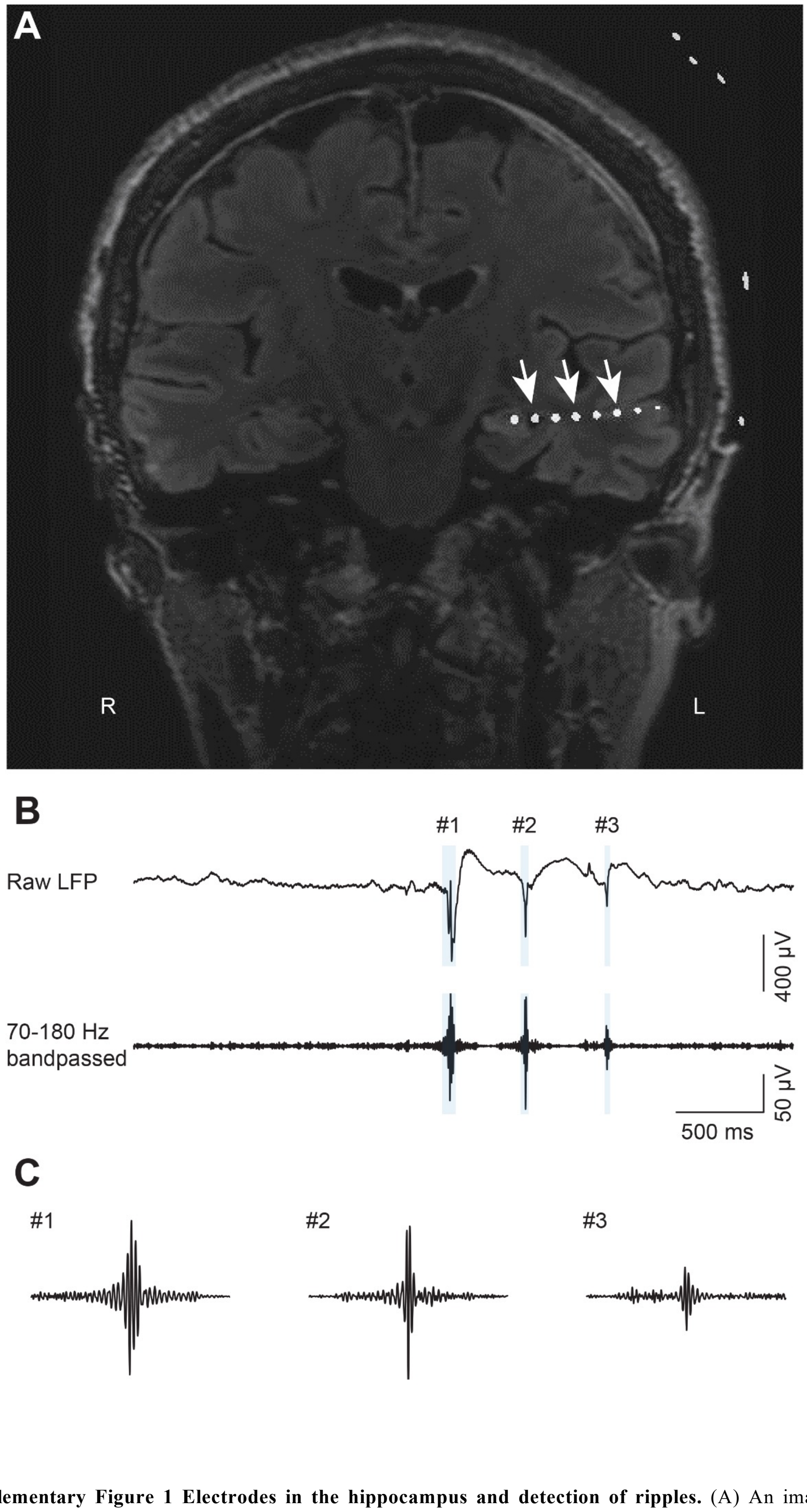


**Supplementary Figure 1 Electrodes in the hippocampus and detection of ripples.** (A) An image combining preoperative magnetic resonance imaging (MRI) and postimplantation computed tomography (CT) data illustrates the placement of electrodes within the hippocampus. (B) Hippocampal LFPs (top) processed using a bandpass filter with a range of 70 to 180 Hz (bottom). The blue shaded area indicates the detected ripples. (C) Three representative SWRs are magnified.

### Figure 2

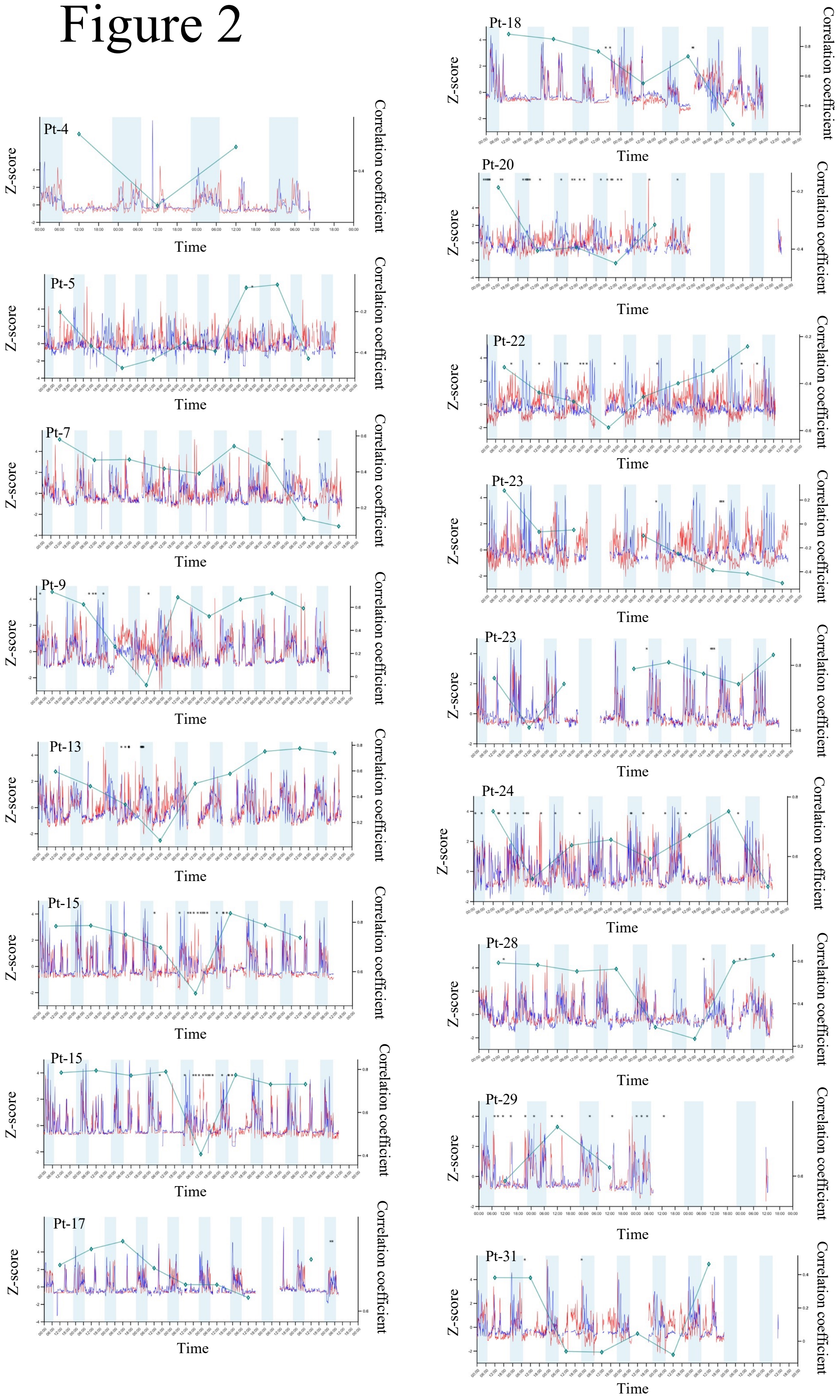


**Supplementary Figure 1 Ripple event rates, delta power and seizures in all cases during recording of iEEG after day 4.** The time courses of the ripple event rates (red) and cortical delta power (blue) in all cases during the recording period after day 4. The light blue background indicates the time of day when the room was darkened. Each square symbol represents the correlation coefficient between the ripple event rate and the delta power for the whole day. Each asterisk indicates the time of a seizure. The x-axis shows the time of day after intracranial electrode implantation, the left y-axis shows the Z-score of ripple event rate or delta band power, and the right y-axis shows the correlation coefficient between the two.

### Figure 3

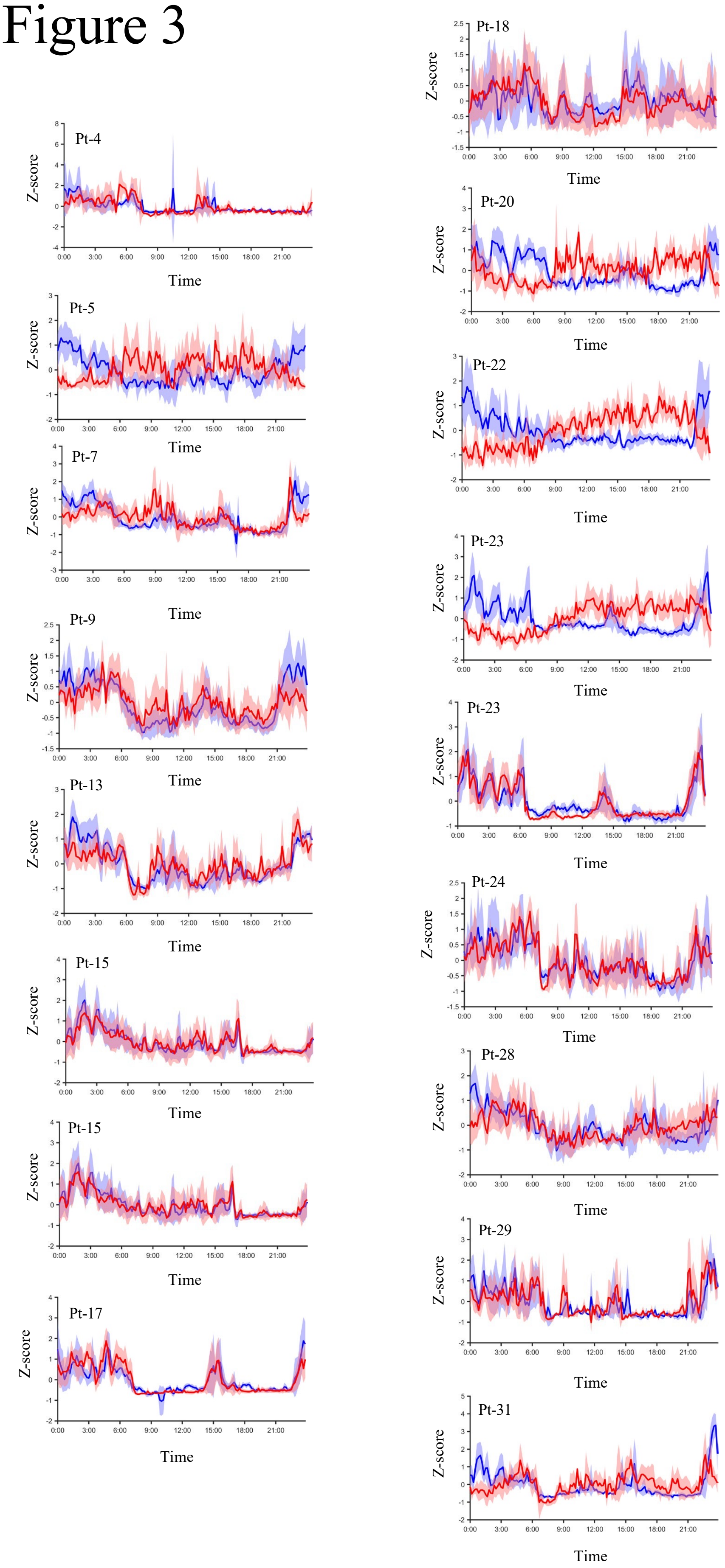


**Supplementary Figure 2 Mean ripple event rates and delta power of each case in iEEG recording after day 4.** The lines show the mean and 95% confidence interval of the Z scored ripple event rates (red) and delta power (blue) over 24 hr for all patients in the NE group and EP group. The x-axis shows time and the y-axis shows the z-score of the ripple event rate or delta band power.

### Figure 4

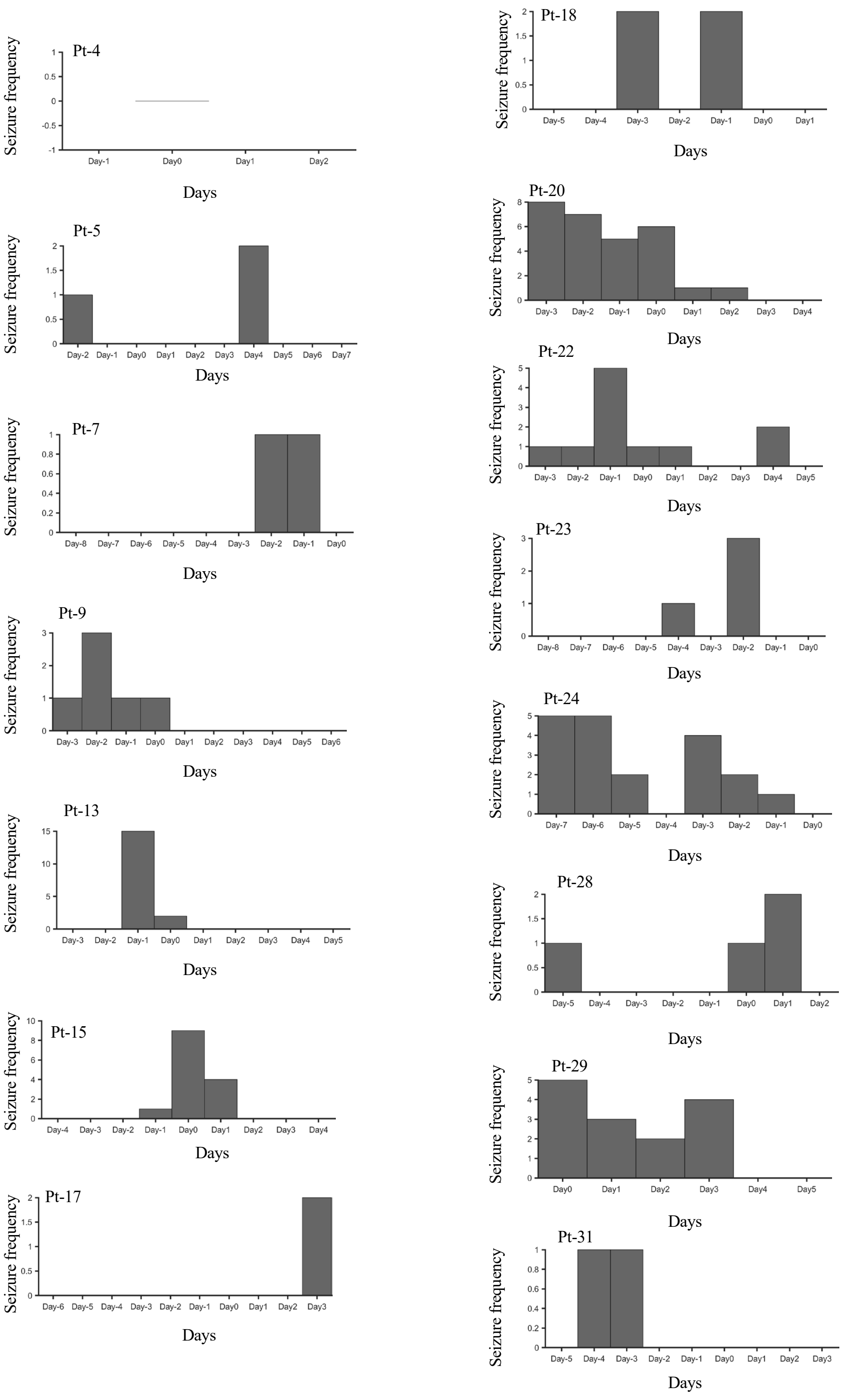


**Supplementary Figure 3 Seizure event frequency around the day with the minimum correlation coefficient.** The seizure event frequency aligned on the day with the minimum correlation coefficient was plotted for all patients in the NE group and EP group. The bar for each day shows the frequency of epileptic seizures for that day. The x-axis shows the days elapsed from the date of the minimum correlation coefficient, and the y-axis shows the frequency of epileptic seizures.

### Figure 5

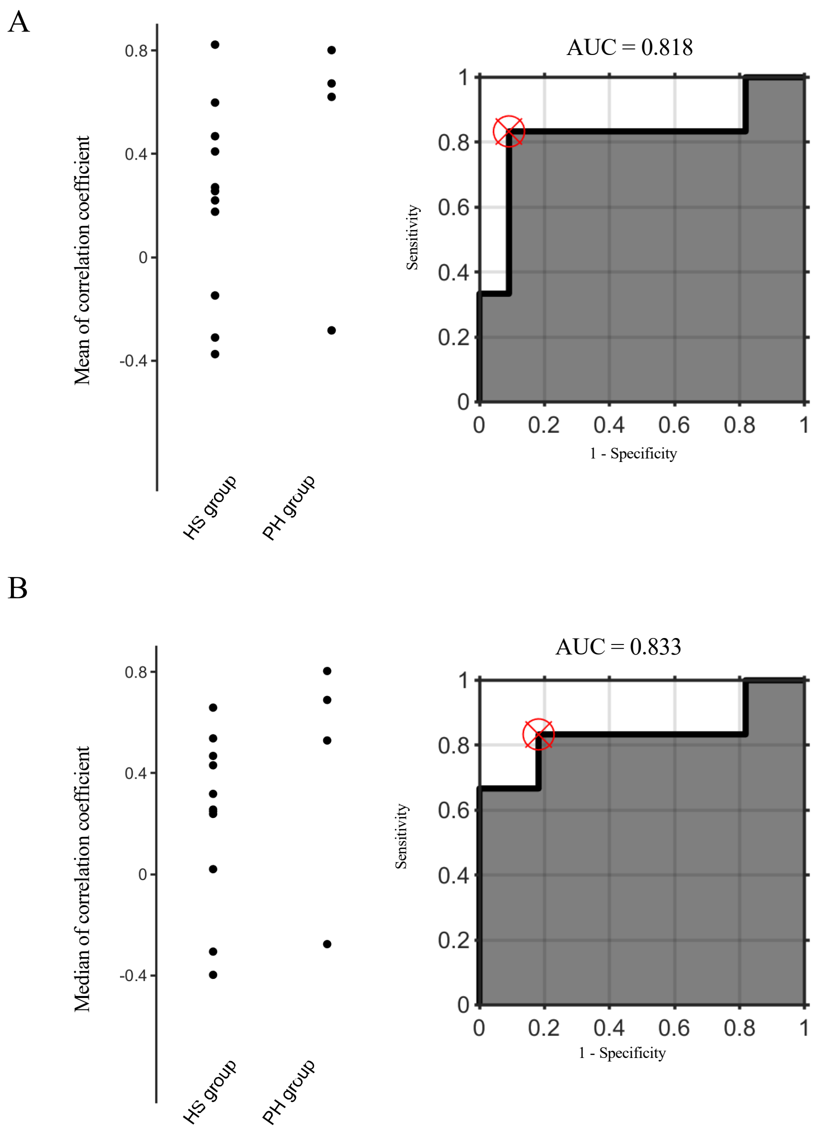


**Supplementary Figure 4 Mean and median value of the correlation coefficient and ROC to classify two groups based on the values.** (**A**) The mean value of the correlation coefficient between the ripple event rates and the delta powers for each patient was plotted for each group (left panel). ROC curve (AUC=0.818) to classify the NE group and EP group based on the mean value of the correlation coefficient (right panel). (**B**) The median value of the correlation coefficient between the ripple event rates and the delta powers for each patient was plotted for each group (left panel). ROC curve (AUC=0.833) to classify the NE group and EP group based on the median value of the correlation coefficient (right panel).

**Supplementary table 1 Information on the amount of anti-epileptic drug for each patient**

| Pt ID | Anti-epilepti c drug | pre ope | Day 1 | Day 2 | Day 3 | Day 4 | Day 5 | Day 6 | Day 7 | Day 8 | Day 9 | Day 10 | Day 11 | Day 12 | Day 13 | Day 14 | Day 15 | Day 16 |
|---|---|---|---|---|---|---|---|---|---|---|---|---|---|---|---|---|---|---|
| 1 | VPA | 1000 | 1000 | → | → | 0 | → | → | → | → | → | → | → | → | → | → | 600 | 1000 |
| | LEV | 2000 | 2000 | 500 | 0 | → | → | 1000 | → | 500 | 0 | 1000 | → | 3000 | → | → | → | → |
| 2 | LEV | 2500 | 2500 | → | → | → | → | → | → | → | → | → | → | → | → | → | | |
| 3 | CBZ | 400 | 400 | → | → | → | 300 | 100 | 0 | → | → | → | → | → | | | | |
| | CLB | 30 | 30 | → | → | → | → | → | → | → | 0 | → | → | → | | | | |
| 4 | CBZ | 200 | 200 | → | → | 100 | 0 | → | 100 | 200 | 200 | → | → | → | | | | |
| | LEV | 1000 | 1000 | 750 | 500 | → | → | 250 | → | 1000 | 1000 | → | → | → | | | | |
| 5 | GBP | 2000 | 2000 | → | → | → | → | → | 1600 | 200 | 0 | → | 800 | 2000 | | | | |
| | TPM | 75 | 75 | → | → | → | → | → | → | → | → | → | → | → | | | | |
| | LEV | 3000 | 1000 | 1500 | 2000 | → | → | → | → | → | → | → | → | → | | | | |
| 6 | PHT | 225 | 225 | 100 | 0 | → | → | → | → | → | → | → | 200 | → | → | | | |
| | CLB | 5 | 5 | → | → | 0 | → | → | → | → | → | → | 5 | → | → | | | |
| | TPM | 300 | 100 | 0 | → | → | → | → | → | → | → | → | 300 | → | → | | | |
| 7 | CLB | 20 | 20 | → | 10 | → | → | → | → | → | → | → | → | 20 | → | | | |
| | LTG | 75 | 0 | → | → | → | → | → | → | → | → | → | → | 75 | → | | | |
| 8 | CLB | 15 | 15 | → | → | → | → | → | → | → | → | → | → | → | | | | |
| | LTG | 350 | 350 | → | → | → | → | → | → | → | → | → | → | → | | | | |
| | LEV | 1500 | 0 | → | → | → | → | → | → | → | → | 750 | 1500 | → | | | | |
| | LCM | 400 | 400 | → | → | → | → | → | → | 200 | 0 | 200 | 400 | → | | | | |
| 9 | CZP | 1 | 1 | → | → | → | → | → | → | → | → | → | → | → | | | | |
| | VPA | 1200 | 1200 | → | → | → | → | → | → | → | → | → | → | → | | | | |
| | GBP | 600 | 0 | → | → | → | → | → | → | → | → | → | → | → | | | | |
| | LEV | 2000 | 1000 | → | → | → | 500 | 0 | → | → | → | 1000 | 2000 | → | | | | |
| | LCM | 100 | 0 | → | → | → | → | → | → | → | → | → | → | → | | | | |
| 10 | PB | 60 | 60 | 60 | 30 | 15 | 0 | → | → | 30 | 60 | | | | | | | |
| 11 | NZP | 5 | 5 | → | → | → | → | → | → | → | → | → | → | → | → | → | → | |
| | CBZ | 400 | 400 | → | → | → | → | → | → | → | → | → | → | → | → | → | → | |
| | LCM | 400 | 400 | 300 | 100 | 0 | → | → | → | 200 | → | 400 | → | → | → | → | → | |
| 12 | PHT | 100 | 100 | → | → | → | → | 50 | 0 | → | → | → | 50 | 100 | → | → | | |
| | ST | 300 | 300 | → | → | → | → | → | → | → | → | → | → | → | → | → | | |
| | CBZ | 600 | 600 | 500 | 400 | → | → | 200 | 0 | → | → | → | → | → | → | 600 | | |
| | LTG | 500 | 500 | → | → | → | → | → | → | → | 475 | 450 | 475 | 500 | → | | | |
| 13 | CBZ | 900 | 600 | → | → | 400 | 200 | → | 900 | → | → | → | → | → | → | | | |
| | VPA | 400 | 400 | → | → | → | → | → | → | → | → | → | → | → | → | | | |
| | ZNS | 50 | 50 | → | → | → | → | → | → | → | → | → | → | → | → | | | |
| | GBP | 400 | 400 | → | → | → | → | → | → | → | → | → | → | → | → | | | |
| 14 | VPA | 1000 | 1000 | → | → | → | → | → | → | 800 | 500 | → | → | → | 700 | | | |
| | LTG | 70 | 70 | → | → | → | → | → | → | → | → | → | → | → | → | | | |
| | LCM | 25 | 25 | → | → | → | → | → | → | → | → | → | → | → | → | | | |
| 15 | CBZ | 400 | 400 | → | → | → | 200 | 100 | 0 | → | 400 | → | → | → | → | | | |
| | LTG | 200 | 200 | → | 150 | 100 | → | → | → | → | 200 | → | → | → | → | | | |
| | PER | 2 | 2 | → | → | → | → | → | → | → | → | → | → | → | → | | | |
| | LCM | 200 | 200 | → | → | → | → | → | → | → | → | → | → | → | → | | | |
| 16 | CBZ | 400 | 400 | → | → | 200 | 0 | → | → | → | → | → | → | → | 400 | | | |
| | LEV | 2000 | 1000 | 0 | → | → | → | → | → | → | → | → | → | → | 2000 | | | |
| | PER | 2 | 2 | → | → | → | → | → | 4 | 2 | → | → | → | → | → | | | |
| 17 | CBZ | 400 | 400 | → | → | → | 0 | → | → | → | → | → | → | → | 400 | | | |
| | LTG | 200 | 200 | → | → | → | → | 0 | → | → | → | → | → | → | 100 | | | |
| 18 | LTG | 210 | 210 | → | → | → | 160 | 60 | → | 110 | → | → | 210 | → | | | | |
| | PER | 6 | 6 | → | → | → | → | → | → | → | → | → | → | → | | | | |
| | LCM | 400 | 400 | 300 | 200 | 100 | 50 | 0 | → | 200 | 400 | → | → | → | | | | |
| 19 | LEV | 1000 | 500 | 1000 | 1000 | 750 | 500 | 750 | 1000 | → | → | → | → | → | → | | | |
| | LTG | 200 | → | → | → | → | → | → | → | → | → | → | → | → | → | | | |
| | LCM | 400 | 0 | → | → | → | → | → | → | → | → | → | → | → | → | | | |
| | CLB | 10 | → | → | → | → | → | → | → | → | → | → | → | → | → | | | |
| 20 | LEV | 1000 | → | → | → | → | → | → | → | → | → | → | → | | | | | |

| | | | | | | | | | | | | | | | | |
|---|---|---|---|---|---|---|---|---|---|---|---|---|---|---|---|---|
| | CBZ | 450 | → | → | → | → | → | → | → | → | → | → | → | | | |
| | VPA | 700 | → | → | → | → | → | → | → | → | → | → | → | | | |
| | ZNS | 300 | → | → | → | → | → | → | → | → | → | → | → | | | |
| | CLB | 30 | → | → | → | → | → | → | → | → | → | → | → | | | |
| 21 | LEV | 2500 | 2000 | → | → | → | 1500 | 1000 | → | → | → | 2500 | → | → | → | |
| | LCM | 250 | → | 200 | → | 250 | 150 | → | → | → | → | 250 | → | → | → | |
| | CLB | 3 | → | → | → | → | → | → | → | → | → | → | → | → | → | |
| 22 | LEV | 1000 | 500 | 1000 | → | → | → | → | → | → | → | → | → | → | → | → |
| | LCM | 300 | 150 | 250 | 200 | 0 | → | → | → | → | → | → | → | → | → | → |
| 23 | LEV | 3000 | 1000 | → | → | → | → | → | → | → | → | → | → | → | → | |
| | PER | 8 | → | → | → | → | → | → | → | → | → | → | → | → | → | |
| | LCM | 200 | → | → | → | → | → | → | → | → | → | → | → | → | → | |
| 24 | LEV | 2000 | 1500 | 2000 | → | → | → | → | → | → | → | → | → | → | → | |
| | PER | 2 | → | → | → | → | → | → | → | → | → | → | → | → | → | |
| | LCM | 100 | 100 | 200 | 200 | → | → | → | → | → | → | → | → | → | → | |
| 25 | LEV | 2500 | → | 1500 | 500 | → | → | → | → | 750 | 1000 | 1500 | 1750 | 2000 | | |
| | PER | 8 | → | → | → | → | → | 2 | → | 4 | 4 | → | 6 | 8 | | |
| | LTG | 400 | → | → | → | → | → | 200 | 100 | 200 | 300 | → | 400 | 400 | | |
| 26 | LTG | 200 | 200 | → | → | 100 | → | → | 50 | → | 75 | 125 | 200 | | | |
| | LCM | 400 | 0 | → | → | → | → | → | → | → | 200 | 400 | → | | | |
| 27 | LEV | 1000 | 750 | 250 | 0 | 750 | 500 | 250 | → | 750 | 500 | 250 | → | → | 0 | 500 |
| | LTG | 125 | 75 | 25 | → | 50 | 25 | → | → | 50 | → | 25 | → | 50 | 0 | 25 |
| 28 | PB | 90 | 90 | → | → | → | → | → | → | → | → | → | → | → | | |
| | VPA | 800 | 800 | → | → | → | → | → | → | → | → | → | → | → | | |
| | LTG | 200 | 200 | → | → | → | → | → | → | → | → | → | → | → | | |
| | LEV | 1500 | 1500 | → | → | → | → | → | → | → | → | → | → | → | | |
| | LCM | 400 | 400 | → | → | → | → | → | → | → | → | → | → | → | | |
| 29 | LCM | 400 | 200 | 200 | 100 | 0 | 100 | 100 | 0 | 200 | 400 | | | | | |
| | VPA | 1000 | 600 | 400 | 400 | 200 | 200 | 300 | 100 | 400 | 800 | | | | | |
| | CLB | 10 | 5 | 5 | 0 | 0 | 0 | 5 | 0 | 0 | 20 | | | | | |
| 30 | LEV | 3000 | 1500 | 0 | → | → | → | → | → | 1500 | 3000 | → | → | → | → | |
| | CBZ | 500 | 500 | → | → | → | → | → | → | → | → | → | → | → | → | |
| 31 | LCM | 200 | | | | | | | | | | | | | | |
| | PHT | 275 | | | | | | | | | | | | | | |
| | CLB | 15 | | | | | | | | | | | | | | |
| 32 | LEV | 2250 | 2250 | 1500 | 1000 | → | 1750 | 2250 | 2250 | → | → | → | → | | | |
| | VPA | 800 | 800 | → | → | → | → | → | → | → | → | → | → | | | |
| | LCM | 400 | 400 | → | → | → | → | → | → | → | → | → | → | | | |
| | PER | 4 | 4 | → | → | → | → | → | → | → | → | → | → | | | |
| 33 | LEV | 500 | 500 | 250 | → | 500 | 250 | 0 | → | → | → | 250 | 500 | → | | |
| | LCM | 400 | 400 | → | → | 300 | 200 | 150 | 100 | → | 200 | 350 | 400 | → | | |

LEV: Levetiracetam, PER: Perampanel, VPA: Valproic Acid, CBZ: Carbamazepine, CLB: Clobazam, GBP: Gabapentin, TPM: Topiramate, PHT: Phenytoin, LTG: Lamotrigine, CZP: Clonazepam, LCM: Lacosamide, PB: Phenobarbital, NZP: Nitrazepam, ST: Sulthiame, ZNS: Zonisamide. The numbers indicated in the antiepileptic drug column represent the daily dosage in milligrams (mg).
